# Tumour region identification guided scoring (TRIGS) and foundation model-based Tumour Infiltrating Lymphocyte scoring are prognostic for pathological complete response/event free survival in the triple negative patients in the PARTNER randomized controlled trial

**DOI:** 10.64898/2026.09.15.26363116

**Authors:** Philip C. Schouten, Melis O. Irfan, Zak Kinsella, Aris Sionakidis, Amy Riddell, Joanna R Worley, Jonny Lay, Sam Casford, Karen Pinilla, Justine Kane, Deborah Whitehorn, Silvia Tarantino, A. Dayimu, N. Demiris, Helena M Earl, Nikola Simidjievski, Elena Provenzano, Jean E Abraham

## Abstract

Assessment of tumour infiltrating lymphocytes (TILs) is a robust prognostic biomarker for HER2-positive and triple negative breast cancer. We aimed to update a previously established pipeline for automated TIL assessment to align to clinical scoring guidelines (tumour region identification guided scoring), foundation model-based lymphocyte detection (SAM-TIL) and compare with alternative methods (HoVerNet, muTILs) and gold standard clinical assessment.

TRIGS and SAM-TIL had an odds ratio of 1.95 (95% confidence interval(ci): 1.22-3.03, p=0.005) and 2.32 (95% ci:1.43-3.77, p=0.001) for predicting pathological complete response (pCR) rate in 166 neoadjuvantly treated patients in the TransNEO cohort. Hazard ratios for overall survival in 277 triple negative and HER2-positive patients in The Cancer Genome Atlas were 0.79 (95% ci: 0.63-1.00, p=0.05) and 0.80 (95% ci: 0.67-0.97, p=0.02) for TRIGS and SAM-TIL. Comparator methods showed similar results. Correlation with gold standard clinical assessment in 285 patients from the PARTNER randomized trial ranged from 0.59-0.69, which is substantially more than interobserver variability for the gold standard. Despite differences with gold standard assessment, no substantial difference in predicting pCR (AUC 0.60-0.64) or event free survival (Integrated Brier score approximately 0.10) were observed between the tested methods and gold standard assessment, suggesting AI tools that do not follow manual scoring guidelines could be validated and subsequently used.

Although we reached prognostic performance similar to published literature and gold standard assessment, the lymphocyte detections produced by the models are not interchangeable with gold standard clinical assessment (correlation 0.59-0.69) and therefore cannot support pathologist assessment. Further studies to validate independent use and/or to improve prognostication are required.

## Introduction

Tumour infiltrating lymphocytes (TILs) have emerged as a robust biomarker with prognostic and predictive implications in triple negative and HER2-positive breast cancer. This biomarker has been incorporated in several guidelines and trials are ongoing to validate the predictiveness (1). It is recommended to count stromal TILs because of reproducibility. sTILs are defined as mononuclear immune cells, lymphocytes and plasma cells, that are resident in the tumour-associated stroma. sTILs are reported as the percentage surface area of TILs in the overall tumour-associated stroma area (2).

The use of machine learning could support faster and reproducible assessment of sTILs and it is recommended that strategies to develop an algorithm strictly adhere to the visual assessment guidelines (1). The current state of the art from a recent review (1) and large scale studies in HER2 positive (3) and triple negative breast cancer (4) demonstrates that manual sTIL scoring is reproducible and that despite automated methods not being interchangeable with manual assessment equivalent or better performance may be obtained. Such automated strategies face practical implications, such as the availability of infrastructure for machine learning, as well as validation in the context of a large body of high-level evidence for following the currently established guidelines. On the contrary, algorithms that have been developed to follow the guidelines encounter other challenges, specifically in model choice and how to translate the manual guidelines into an executable approach, that preclude their widespread use without further validation (1). Additionally, during this study, ESMO published guidelines on the implementation and validation of AI algorithms in oncology, and for this study the Class A (AI for biomarker qualification) and Class C1 (novel AI based prognostic biomarkers) pathways are of relevance. The former requires a concordance study comparing to gold standard, the latter retrospective analysis of high-quality clinical trial data comparing to validated prognostic markers (manual sTIL) and clinical endpoints (pCR and survival)

The gaps this study addresses are threefold and ultimately aim for implementation of automated support or assessment of pathology biomarkers in a real-world setting of prospective clinical trials. 1) Update an in-house model (baseline) with detection of tumour regions to match manual assessment. Lymphocyte density was previously established as a predictor of response to neoadjuvant chemotherapy (5) and validated in a retrospective analysis of an independent randomized trial (6) and included in an integrated model of clinical, pathological and genetic predictors of response (7). The TIL density algorithm comprised a support vector machine to classify cells as lymphocyte, tumour and stromal, and did not include identification of relevant regions. With the advent of current deep learning approaches for computer vision, we envisaged that the existing pipeline could be improved by implementing a more robust cell classification. Additionally, we sought to implement and validate changes that make the pipeline adhere better to the clinical manual scoring guidelines by first identifying regions of interest to count lymphocytes in. 2) Foundation models hold a promise to supply a stable base architecture and performance on a wide variety of tasks (8,9). During the study few foundation model methods became available other than our approach of using Segment Anything for pathology (8), including prompting a vision language model which resulted in suboptimal performance (10), methods that regress on foundational model embeddings, which do not follow the manual approach (11,12) and CellVit++, where Segment anything for histopathology had as benefits outperformance on overall nuclei segmentation, being large and not huge and possibility to use interactively with QuPath (13). 3) Robust evaluation of various automated methods and assessment of performance within a clinical workflow with the aim to select a suitable algorithm.

## Methods

### Datasets

TransNEO – The TransNEO neoadjuvant breast cancer molecular profiling study (7) contains 204 whole slide images scanned at 20X on an Aperio scanner of frozen pre-treatment biopsies from 168 patients from a neo-adjuvant cohort comprising both hormone receptor positive and negative and HER2 positive and negative cases. 166 of the patients contain an assessment of patient outcomes e.g. residual disease or pathological complete response. Whole slide image (WSI) were available at https://zenodo.org/records/6337925#.Y30d1y-l1Ls and clinical data and methods through the previous publication (7).

TCGA - The TCGA dataset contains WSIs and clinical data of 276 patients from the HER2-positive and triple negative breast cancer cohort with complete survival data extracted from the Genomics Data Commons Portal (https://portal.gdc.cancer.gov/) using the TCGAlinks Bioconductor R package.

PARTNER - The PARTNER dataset contains pre-treatment formalin fixed paraffin embedded biopsies of 285 patients were scanned at 20X at an Aperio scanner. Clinical data was obtained through the PARTNER randomized clinical trial (14,15). The selection of this subset was done based on tissue availability for whole genome sequencing. The PARTNER trial protocol (NCT03150576) was approved by the North West – Haydock Research Ethics Committee (ref: 15/NW/0926) and the trial was carried out in accordance with the Declaration of Helsinki and the European Clinical Trials Directives 2001/20/EC.

### Computational pipeline/analysis

Support Vector Machine (SVM) - Identification of lymphocytes within the TransNEO digital slides was achieved with an SVM supervised machine learning algorithm, which identifies tissue and cells and classifies cells as lymphocytes, cancer or stroma (5). Lymphocyte counts and densities were made available as part of the TransNEO data release enabling the comparison of our TransNEO TIL densities to those calculated in the previous publication (7).

HoVerNet - Hover-Net is a convolutional neural network (CNN) which uses spatial information along the horizontal and vertical axes of a cell to identify and classify the cell type (16). We use a version of HoVer-Net which has been trained on MoNuSAC Grand Challenge data (17) as it specifically includes lymphocytes as a class. In this implementation the whole WSI is annotated, and the other output classes (specifically the epithelial class) are not used. We use the TIAToolbox (18) repository to implement the MoNuSAC trained Hover-Net.

Tumour Region Identification Guided Scoring (TRIGS) - To identify lymphocytes in and in proximity of the tumour, we extend the functionality of the MoNuSAC Hover-Net segmentation by taking a two-stage approach to segmentation. First, we use the semantic segmentation functionality of TIAToolbox to identify tumour regions through the use of a CNN trained on the Breast Cancer Semantic Segmentation (BCSS) dataset (19). After semantic segmentation we expand the tumour masks at a lower resolution than the full image resolution and apply a wavelet transform (20) to include the surrounding stroma. Secondly, we apply the MoNuSAC trained Hover-Net to identify lymphocytes only within the regions of the tissue sample which are highlighted by the tumour mask (just as in the HoVerNet implementation we do not use the epithelial output class). In practise any combination of semantic and single cell segmentation algorithm could be employed to achieve a measurement of TIL density; the two-stage nature of the methodology is the key feature as opposed to a specific algorithm. We refer to this two-stage pipeline as Tumour Region Identification Guided Scoring (TRIGS).

SAM-TIL - ‘Segment Anything’ is a model based on the Vision Transformer (ViT) architecture, trained on 1 billion masks over 11 million images for segmentation of natural images (21). We used a version of this model that has been trained on public microscopy (22), and pan-cancer histopathology datasets (8). We used previously described methods (8) to fine-tune this histopathology-directed variant (ViT-L-histopathology) of SAM for semantic segmentation of output classes ‘Background’, ‘Epithelium’, ‘Stroma’, ‘TIL’, using a public 40x magnification TIL dataset (23).

Briefly, we trained a UNETR decoder (24) from scratch using the output of the frozen ViT-L-histopathology image encoder, optimized by a weighted cross-entropy loss for the frequency of classes in the entire dataset. Train-test was split randomly 80:20. We used an AdamW optimizer used with an initial learning rate of 10-4, implementing a learning rate scheduler with a factor of 0.9 and patience of 5 epochs. The model was trained for 100,000 iterations. The epoch with the best validation metric was chosen for downstream use.

During inference, we generate two outputs: instance and semantic segmentation. Instance segmentation was generated by the ViT-L-histopathology model as this is the current best-in-class model for automatic instance segmentation in histopathology images. Semantic segmentation was generated by our TIL-finetuned model. To unify outputs into a single representation, we generate panoptic segmentation masks (23) by encoding each pixel’s semantic label and instance ID into a unique integer, using an offset of 10,000,000 to accommodate large frequencies of detected cell instances over WSIs. All panoptic operations were vectorized using NumPy. For processing TCGA images, we used WSInfer (25) to generate tissue/tumour masks that were passed to the SAM-TIL pipeline.

MuTILS - muTILs utilises a two-stage identification approach; first semantic segmentation performed by a CNN highlights regions within the tissue sample of clinical interest and then single cell segmentation performed by a separately trained CNN identifies lymphocytes within these regions (23). To our knowledge this algorithm best resembles visual scoring guidelines.

### Statistical analysis

Descriptive patient characteristics were obtained as continuous or categorical values. Spearman’s rank correlation coefficients were calculated between the TIL densities obtained from the various methods. (Multivariable) logistic regression and Cox regression analyses were performed to predict pathological complete response and event free survival respectively in the TransNEO and PARTNER datasets. A comparison between the gold standard clinical method, the automated analysis most resembling clinical guidelines and our newly trained methods were performed in the PARTNER trial data, using multiple methods: Harrell’s concordance index (26), Brier score (27,28), Cumulative/Dynamic (C/D) AUC using the estimator proposed by Uno et. al (29), Integrated Brier score, overall measure of survival model performance across all time points, Integrated C/D AUC, overall measure of survival model performance across all time points. Calculations were done in R and/or Python version 3.9, scikit-learn version 1.5.2, statsmodels version 0.14.6 and lifelines version 0.30.0. Code will be made available where appropriate and can be requested from the authors by reasonable request.

## Results

### Analysis of the TransNEO dataset

We used the TransNEO set as a training set, as it had previously established TIL density scores, which allowed us to compare HoVerNet implementation with the previously validated SVM. Figure 1 shows a representative high magnification HCE, density plots across a WSI for the various methods. Figure 2 shows correlation between the various methods. Semi-quantitative assessment (PS) of x slides and correlation coefficients show reasonable to good concordance between the methods.

**Figure 1.**
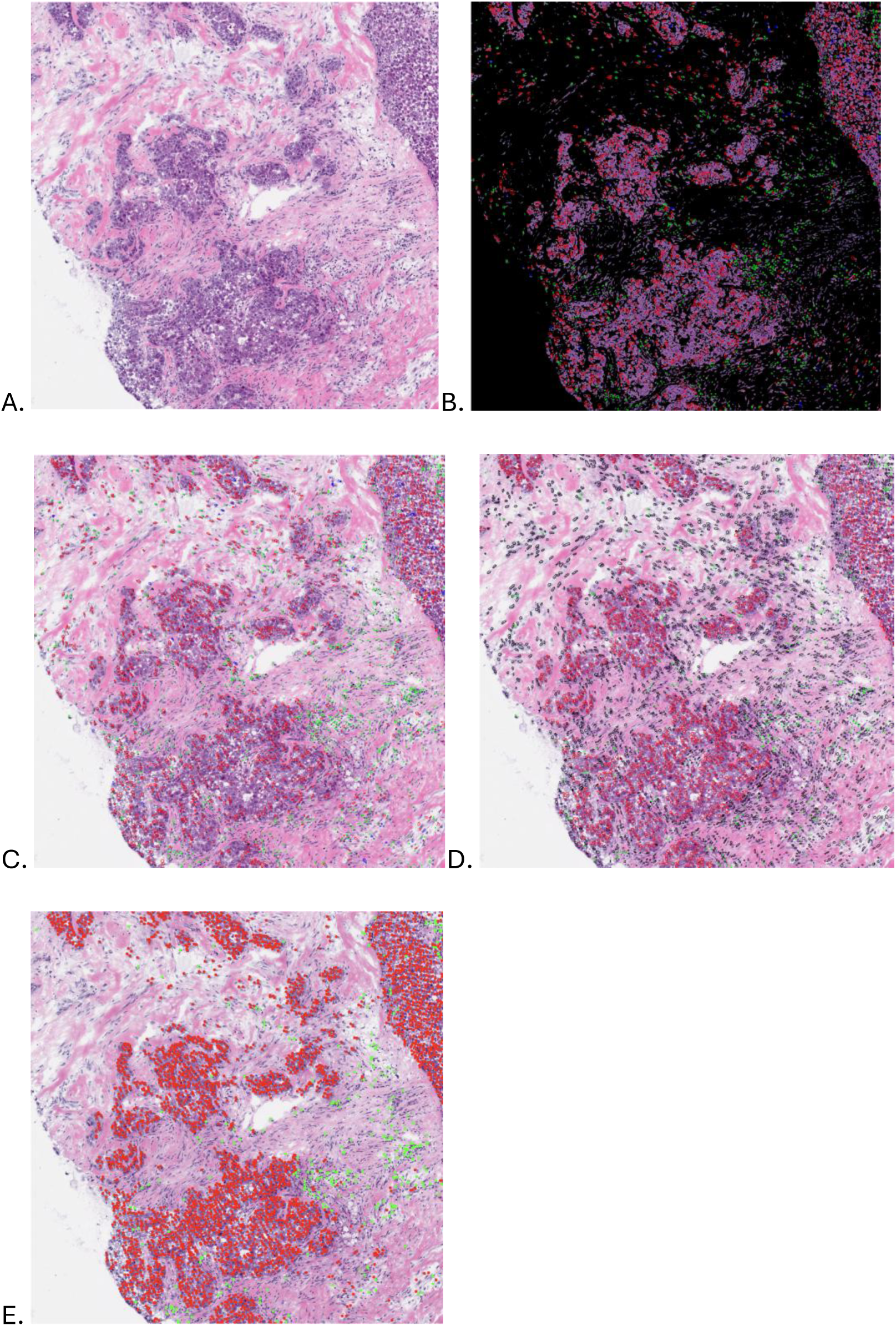
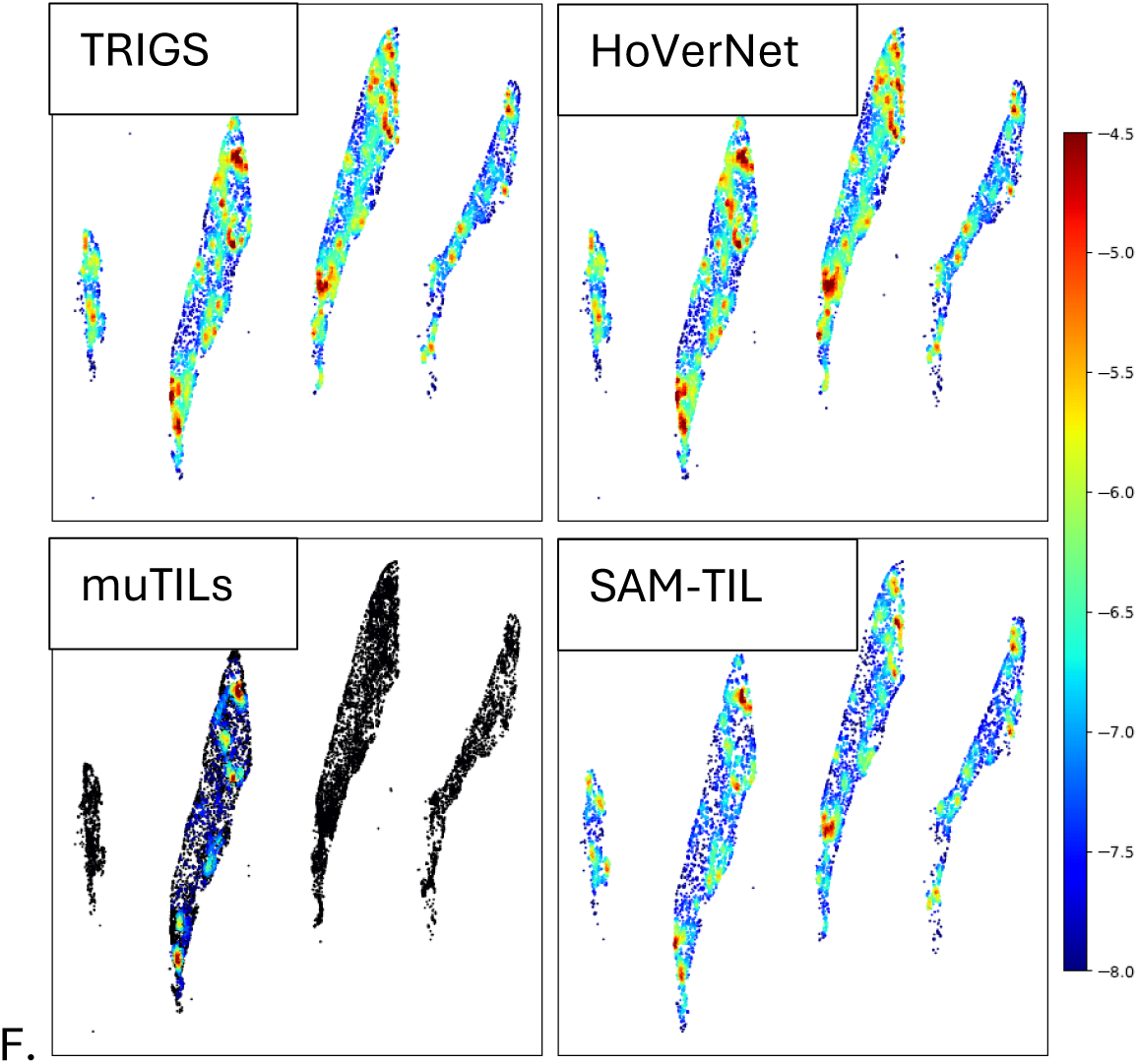
Example tile with annotations of the tested methods. A. HCE staining. B. TRIGS segmented slide, black: masked, red: epithelial, green: lymphocytes. C. HoVerNet segmented slide, red: epithelial, green: lymphocytes. D. SAM-TIL segmented slide, red: neoplastic, green: lymphocytes, black stromal. E. MuTILs segmented slide, red: cancer epithelial, green: lymphocytes. F. Heatmap of TIL density across a whole slide image, colour scale represents the density, with higher densities shown in red. Black areas are excluded from the calculation.

**Figure 2.**
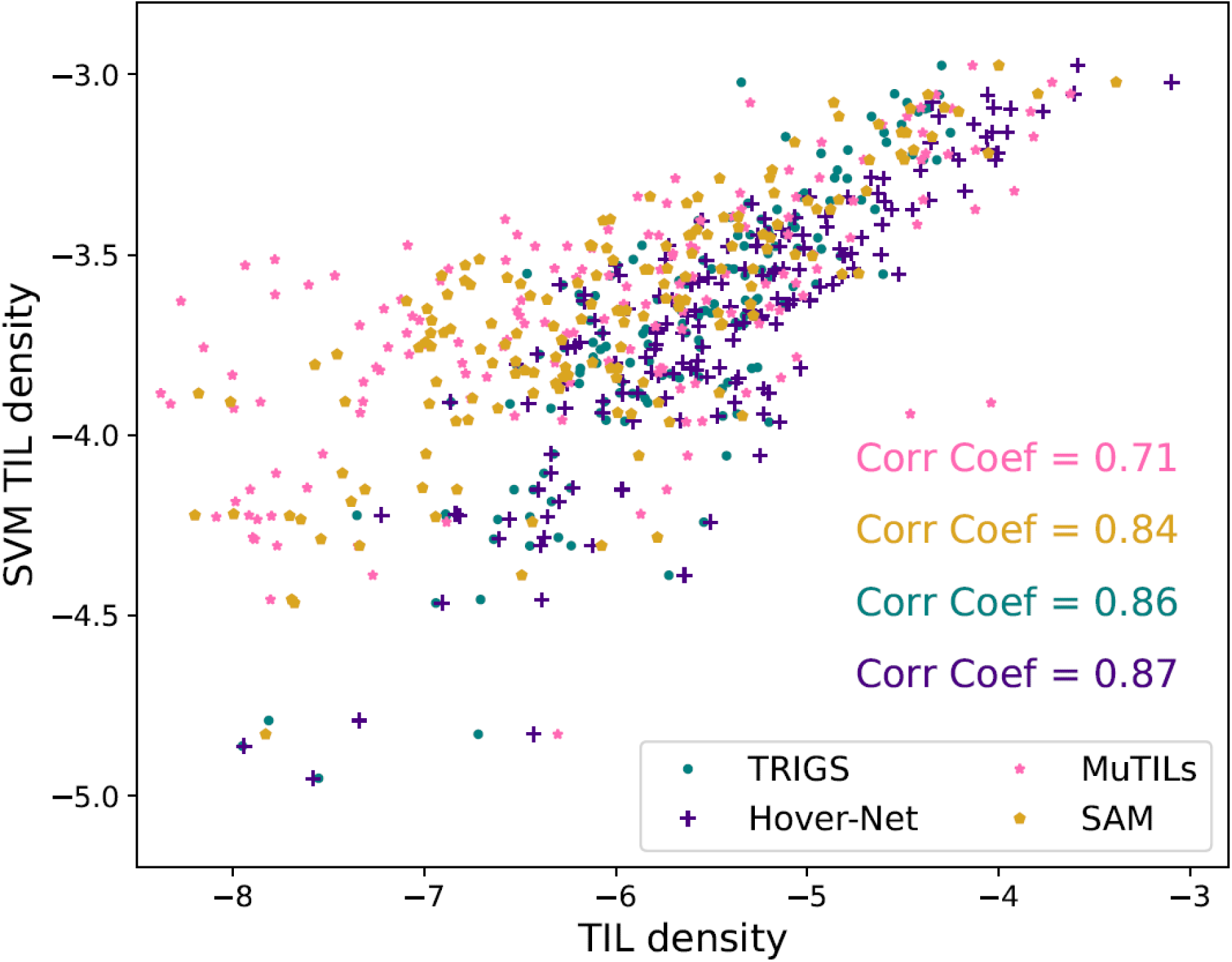
Correlation between the median TIL density per TransNEO slide as calculated by various pipelines and the SVM.

Subsequently, we performed logistic regression analysis to assess whether the previously described correlation of the SVM with pathological complete response status was present for the new and comparator methods (Table 1). Out the total 204 TransNEO slides, digital pathology results for the SVM method were made publicly available for 166 slides. HoVerNet, TRIGS, SAM-TIL produced TIL counts for 166 slides, while MuTILs produced TIL counts for 164 slides. There is substantial overlap between odds ratios and confidence intervals, with perhaps muTILs being on the lower end of the spectrum. Of note is that muTILs substantially differs in the proportion of the slide selected for analysis (not covering either the whole slide, such as the SVM and SAM TIL, and not covering all tumour such as TRIGS). In addition, muTILs was not trained on frozen tissue slides.

**Table 1.** Logistic regression analysis of TransNEO.

|  |  | SVM |  |  | TRIGS |  |  | HoVerNet |  |  | muTILs |  |  | SAM |  |  |
| --- | --- | --- | --- | --- | --- | --- | --- | --- | --- | --- | --- | --- | --- | --- | --- | --- |
| Variable | Value | OR | CI | p | OR | CI | p | OR | CI | p | OR | CI | p | OR | CI | p |
| Node | > or <= 50 mm | 1.65 | 0.70-3.87 | 0.250 | 1.70 | 0.73-3.93 | 0.217 | 1.62 | 0.69-3.78 | 0.265 | 1.67 | 0.73-3.83 | 0.223 | 1.55 | 0.66-3.66 | 0.317 |
| Grade | 1,2 or 3 | 2.07 | 0.76-5.63 | 0.155 | 2.12 | 0.80-5.63 | 0.133 | 2.12 | 0.78-5.76 | 0.139 | 2.29 | 0.87-6.00 | 0.091 | 2.12 | 0.88-5.08 | 0.145 |
| ER | 0 or 1 | 3.64 | 1.50-8.83 | 0.004 | 3.63 | 1.52-8.68 | 0.004 | 3.53 | 1.46-8.49 | 0.005 | 3.21 | 1.37-7.50 | 0.007 | 3.89 | 1.59-9.49 | 0.003 |
| HER2 | 0 or 1 | 2.12 | 0.88-5.10 | 0.094 | 2.16 | 0.91-5.15 | 0.081 | 1.99 | 0.83-4.73 | 0.121 | 1.75 | 0.75-4.06 | 0.196 | 2.11 | 0.88-5.08 | 0.094 |
| TIL | continuous | 2.46 | 1.41-4.30 | 0.002 | 1.93 | 1.22-3.03 | 0.005 | 2.10 | 1.33-3.33 | 0.002 | 1.50 | 0.97-2.31 | 0.065 | 2.32 | 1.43-3.77 | 0.001 |

### TCGA Analysis

We selected all HER2-positive and triple negative cases from TCGA and applied our methods to WSI of 276 patients. Out of the 293 relevant TCGA slides HoVerNet processed 280 slides, TRIGS processed 291 slides, SAM-TIL processed 287 slides and MuTILs processed 283 slides. Like TransNEO there is substantial concordance in correlation between methods. Table 2 shows the multivariable Cox Proportional Hazards analysis shows overlapping hazard ratios/confidence intervals and independent prognostic value of TILs obtained with all methods.

**Table 2.** Multivariable Cox Proportional Hazards modelling of overall survival and TIL scores.

|  |  | TRIGS |  |  | HoVerNet |  |  | muTILs |  |  | SAM |  |  |
| --- | --- | --- | --- | --- | --- | --- | --- | --- | --- | --- | --- | --- | --- |
| Variable | Value | HR | CI | p | HR | CI | p | HR | CI | p | HR | CI | p |
| ER | 0 or 1 | 0.73 | 0.28-1.92 | 0.53 | 0.81 | 0.29-2.31 | 0.69 | 0.71 | 0.27-1.86 | 0.48 | 0.67 | 0.26-1.76 | 0.42 |
| HER2 | 0 or 1 | 1.17 | 0.46-2.99 | 0.74 | 1.02 | 0.38-2.79 | 0.96 | 1.26 | 0.49-3.23 | 0.63 | 1.22 | 0.48-3.12 | 0.67 |
| TIL | continuous | 0.79 | 0.63-1.00 | 0.05 | 0.79 | 0.63-0.99 | 0.04 | 0.81 | 0.68-0.96 | 0.02 | 0.81 | 0.67-0.99 | 0.04 |

### PARTNER Analysis

The PARTNER dataset served as independent test set, as it comprised our highest level of evidence dataset, and included ground truth histopathologist scores. We applied our methods to 285 pre-treatment biopsy specimen WSIs. Figure 3 shows correlation between methods and gold standard pathologist assessment, it is noted that variability is substantial higher than the < 5% interobserver variability aimed for clinically. Subsequently, we used ROC curve analysis and multivariable logistic regression for predicting pathological complete response rate and multivariable Cox proportional hazard modelling to predict Event Free Survival.

**Figure 3.**
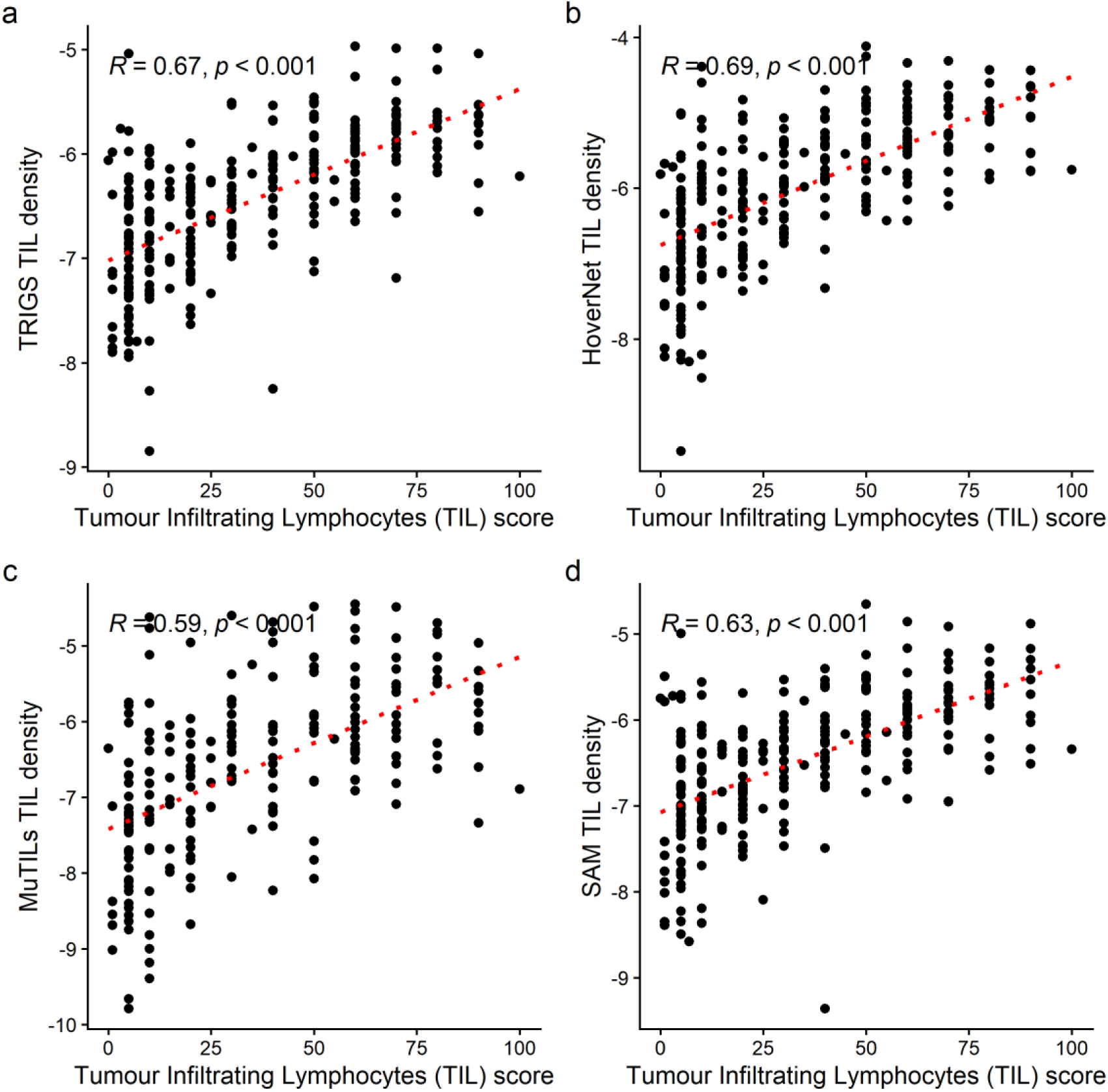
Automated assessment vs. pathologist assessment of TILs. A) TRIGS vs. pathologist. B) HoVerNet vs. pathologist. C) MuTILs vs. pathologist. D). SAM TIL vs pathologist.

**Figure 4.**
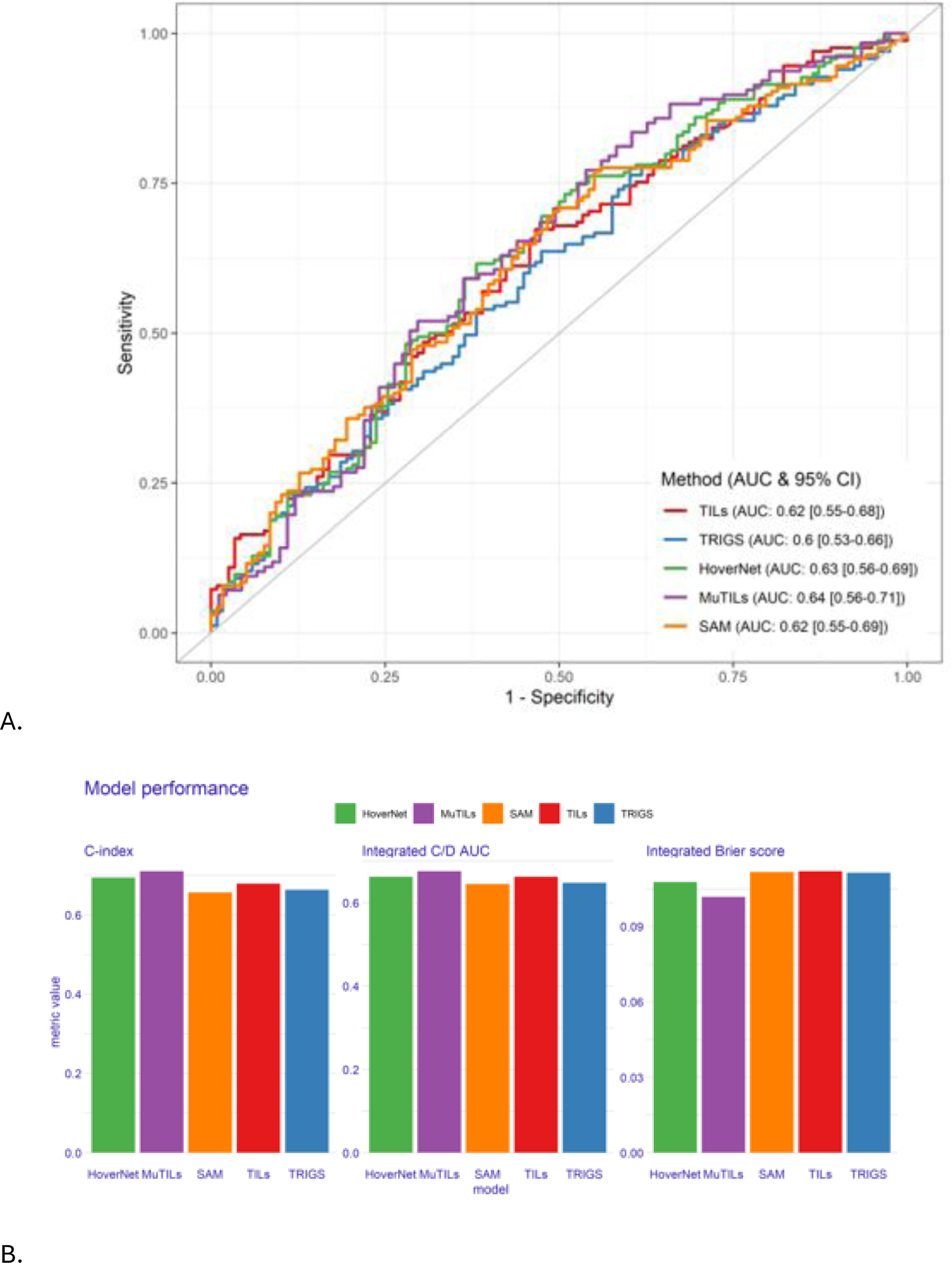
A. Receiver Operating Characteristic (ROC) curve for five TILs scoring method to predict pCR after adjusting for covariates. The x-axis represents the False Positive Rate (1 - Specificity), and the y-axis represents the True Positive Rate (Sensitivity). The Area Under the Curve (AUC) for the five method was similar, indicating all performs similarly. B. C-index, Integrated Brier score and cumulative/dynamic (C/D) AUC of five TILs scoring method.

### Qualitative/semi-quantitative analysis of all datasets

Out of the 285 PARTNER slides HoVerNet processed 284 slides, TRIGS and SAM-TIL processed 285 slides, MuTILs processed 220 slides. Semi-quantitative/qualitative analysis of divergent slides of TransNEO and PARTNER and a wider range of slides from PARTNER was done by PS. Whereas identification of TILs was generally reliable, presumably due to their distinctive cytology, false positives/negatives were seen, e.g. cells cut in wrong plane, debris, small round tumour cells, and benign epithelial nuclei. Unexpectedly, some divergence between methods was observed, for HoverNet a proportion of slides with weaker staining/scanning failed to detect substantial parts of tissue. muTILs failed to detect relevant tumour region for some of the slides. Assessment was limited due to practical constraints, it is beyond the scope of this study to improve on or validate underlying methods, such as tumour detection or lymphocyte detection.

## Discussion

In this study we did a robust evaluation of existing and newly trained methods to estimate/count tumour infiltrating lymphocytes and demonstrated state of the art performance compared to other methods and with similar performance as pathologist estimates using the gold standard guidelines.

This study highlights challenges of implementing and validating AI in clinical trial practice. Measuring tumour infiltrating lymphocytes is arguably a low-complexity task. From a histological perspective, cytology of lymphocytes is uniform and characteristic. In terms of availability of tools, pre-trained neural networks and pipelines are available that provide basic functionality to address sTIL measurements.

We set out to build tools that follow the clinical histopathological guidelines, requiring identification of tumour regions and capturing the surrounding stroma and identifying the lymphocytes in those regions (2). Using TIAToolBox we employed BCSS-pretrained UNET with ResNet50 encoder tumour segmentator and MonuSac pretrained lymphocyte HoVerNet segmentation and classification to identify the relevant region of the slides and the number of lymphocytes (TRIGS). A separate approach finetuned a histopathology-directed large vision model to identify lymphocytes across whole slide images (SAM-TIL). Notably, across datasets we did not observe substantial difference in correlation or prediction of clinical outcomes for methods that pre-select tumour regions (TRIGS, muTILs) vs. comparator methods that did not pre-select (SVM, HoVerNet and SAM-TIL). For datasets containing biopsy specimens (TransNEO and PARTNER) this may reflect selection of most relevant slide by pathologist, who tends to include the slide with most tumour for scanning. However, for the TCGA data which contained larger excision specimens whole slide assessment with HoVerNet did not perform different than the other methods which preselected tumour regions (TRIGS, muTILs and also SAM-TIL for computational efficiency/cost reduction).

Using a method that follows the clinical guidelines, allows for readily knowing the expected performance and validating of individual parts, such as tumour detection or lymphocyte classification. When better tools become available, these components can be replaced in a modular fashion. However, such approach comes also with a burden of running multiple networks and for example in the TCGA data we encountered efficiency issues with implementation and large amount of computation to cover the whole slide. Recently, a weakly supervised regression from a histopathology-trained feature extractor was proposed (11). This method does not directly follow the same instructions but reaches good correlation with pathologist scores. Unwanted/unexpected behaviour such as identifying invasive lobular cancer as lymphocytes, and lymphovascular invasion, immune hotspots, fibrotic areas and lymphocytes at the tumour boundary which would not be counted by pathologists are similar for methods we proposed and validated against.

Although following clinical guidelines supports explainability, it may not leverage AI’s capability of actually counting TILs on the slide, something that is inherently impossible within the time constraints of normal clinical practice. Other approaches than adhering to the guidelines could be equally explainable (i.e. showing location and amount of TILs identified by the method), or able to be validated by calibrating against pathologist scores or another gold standard, like immunohistochemistry. Given the extent of literature showing that many different approaches show prognostic value (1), it may be that TILs are such a robust biomarker that the method matters less (even if certain region selections/segmentations or cell classifications are verifiably wrong), which has also been suggested by Vidal et al (30). Alternatively, it may be that the series we have investigated are too small to show differential effect between the methods; if that were the case, the differences may not be clinically meaningful. In the meantime, adherence to clinical guidelines and pathologist overview of automated TIL assessment would be recommended, with algorithmic assessment as possible support but not to independently used.

In the research clinical setting, slides will be available through various labs/protocols and scanners. In this study we have not specifically addressed variability that has been observed (31), and note that there is some robustness against this variability, including that muTILs which was trained on FFPE samples still was prognostic when analyzing frozen biopsy specimens. However, we also noted that muTILs did not produce outcomes for a smaller proportion of PARTNER slides and that basic HoVerNet implementation was subject to issues with faded slides.

Parts of this study were designed before availability of the ESMO guidelines (32). These guidelines form a useful tool for study design, as in the current manuscript we have followed two paths, aiming to stay close to clinical guidelines (biomarker class A: AI for biomarker quantification) but also to compare with other methods and clinical routine gold standard (class C1: novel prognostic biomarker). In our comparison we show correlation coefficients of between 0.59 and 0.69 with the gold standard, which falls short of the high concordance with clinical standard as discussed in the ESMO guideline, because clinically an interobserver variability at <5% is aimed for (11). Interestingly, as observed above, given the C1 biomarker results, alternative approaches may be viable, attractive from a perspective of computational resources and may potentially leverage AI to do tasks that are unavailable for manual assessment, if and when properly validated.

Concluding, we trained and validated methods for automated TIL assessment, with similar performance as gold standard and comparator methods. The methods are substantially different from the gold standard and therefore require extensive validation beyond demonstrating similar prognostic performance. The methods we investigate demonstrate behaviour that require pathologist supervision for real world use. Validation in future studies and comparison with other methods, removal of unwanted behaviour (e.g. by improving underlying components), or assessment as ‘new’ prognostic biomarker are directions for future research.

## Data Availability

All data and code produced in the present study are available upon reasonable request to the authors

## Acknowledgements

This work was supported by the Cancer Research UK Cambridge Centre [CTRQQR-2021\100012]

The Human Research Tissue Bank is supported by the NIHR Cambridge Biomedical Research Centre (NIHR203312)

The PARTNER study was funding by AstraZeneca and CRUK

The United States Department of Defence, FY22 Breast Cancer Research Program of the Congressionally Directed Medical Research Programs, Clinical Research Extension Award GRANT1376971 (Synergia, which includes samples from PARTNER and TransNEO)

## Supplemental Data

**Supplementary Table 1.**
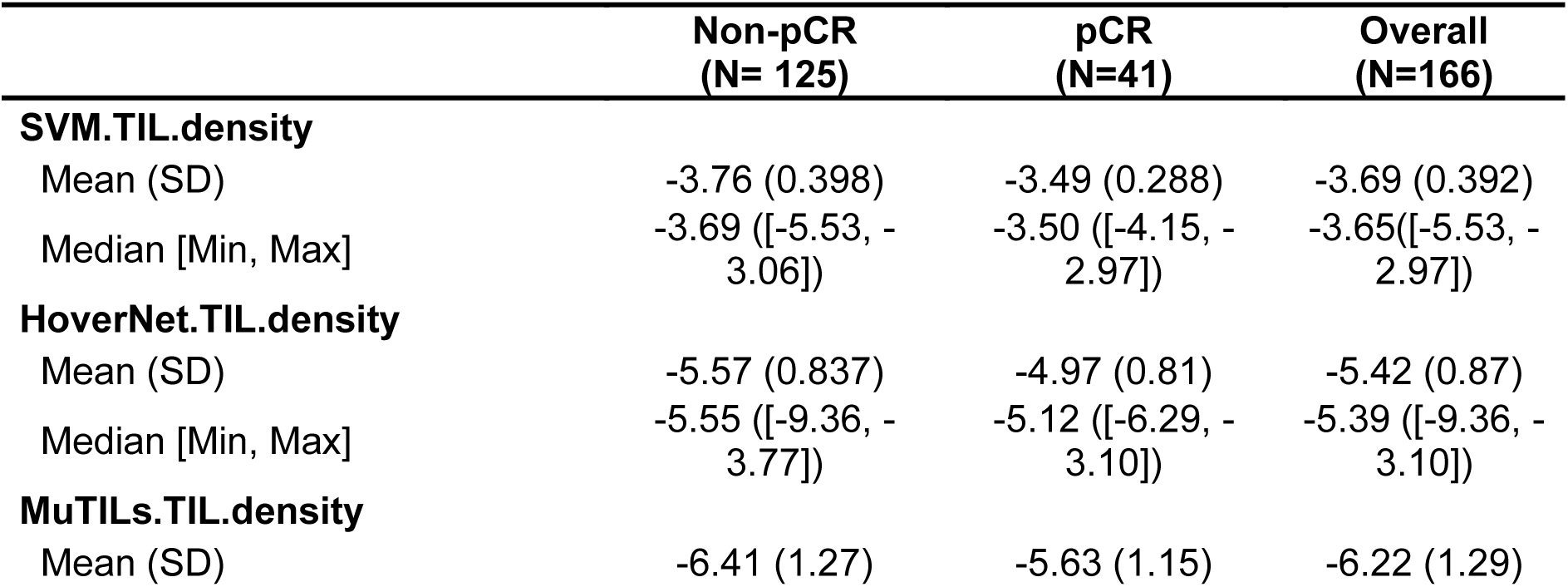
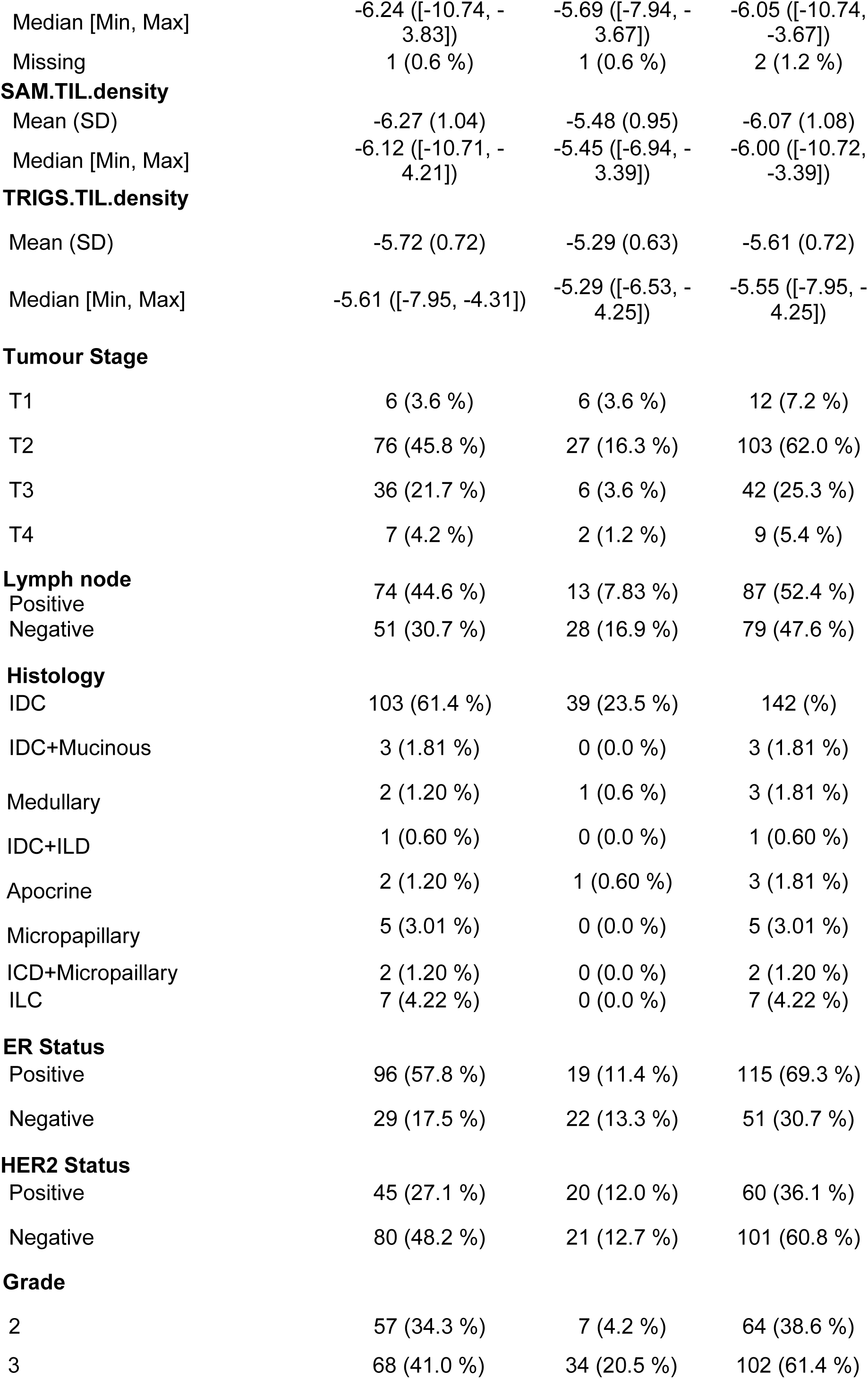
TransNeo.

**Supplementary Table 2.** TCGA.

|  | < 698 days<br>(N= 138) | > 698 days<br>(N=138) | Overall<br>(N=276) |
| --- | --- | --- | --- |
| **SAM.TIL.density** |  |  |  |
| Mean (SD) | -6.80 (1.52) | -6.69 (1.45) | -6.74 (1.51) |
| Median [Min, Max] | -6.67 ([-11.20, -3.73]) | -6.46 ([-12.19, -4.54]) | -6.49 ([-12.19, -3.73]) |
| Missing | 2 (0.72 %) | 4 (1.45 %) | 6 (2.17 %) |
| **HoverNet.TIL.density** |  |  |  |
| Mean (SD) | -6.80 (1.32) | -6.76 (1.21) | -6.76 (1.26) |
| Median [Min, Max] | -6.61 ([-10.67, -3.86]) | -6.65 ([-11.42, -4.53]) | -6.59 ([-11.42, -3.86]) |
| Missing | 6 (2.17 %) | 7 (2.54 %) | 13 (4.87 %) |
| **MuTILs.TIL.density** |  |  |  |
| Mean (SD) | -7.17 (1.58) | -7.13 (1.45) | 7.15 (1.52) |
| Median [Min, Max] | -6.97 ([-10.88, -3.75]) | -6.95 ([-12.70, -4.66]) | -6.96 ([-12.70, -3.75]) |
| Missing | 3 (1.09 %) | 7 (2.62 %) | 10 (3.63 %) |
| **TRIGS.TIL.density** |  |  |  |
| Mean (SD) | -6.97 (1.24) | -6.91 (1.22) | -6.92 (1.22) |
| Median [Min, Max] | -6.82 ([-10.84, -4.05]) | -6.73 ([-11.78, -5.04]) | -6.74 ([-11.78, -4.05]) |
| Missing | 0 (0 %) | 2 (0.72 %) | 2 |
| **Stage** |  |  |  |
| I | 13 (4.71 %) | 24 (8.70 %) | 37 (13.41 %) |
| II | 82 (29.7 %) | 86 (31.16 %) | 168 (60.87 %) |
| III | 36 (13.04 %) | 25 (9.06 %) | 61 (22.10 %) |
| IV | 3 (1.09 %) | 2 (0.72 %) | 5 (1.81 %) |
| X | 1 (0.36 %) | 0 (0 %) | 1 (0.36 %) |
| Unknown | 3 (1.09 %) | 1 (0.36 %) | 4 (1.45 %) |
| **Age** |  |  |  |
| ≤ 60 | 71 (25.72 %) | 93 (33.70 %) | 164 (59.42 %) |
| > 60 | 66 (23.91 %) | 46 (16.67 %) | 112 (40.58 %) |
| **Histology** |  |  |  |
| Infiltrating Lobular Carcinoma | 19 (6.88 %) | 13 (4.71 %) | 32 (11.59 %) |
| Infiltrating Ductal Carcinoma | 113 (40.94 %) | 109 (39.50 %) | 222 (80.43 %) |
| Other | 2 (0.72 %) | 6 (2.17 %) | 8 (2.90 %) |
| Mixed | 0 (0 %) | 6 (2.17 %) | 6 (2.17 %) |
| Metaplastic Carcinoma | 4 (1.45 %) | 2 (0.72 %) | 6 (2.17 %) |
| Medullary Carcinoma | 0 (0 %) | 2 (0.72 %) | 2 (0.72 %) |
| **ER Status** |  |  |  |
| Positive | 61 (22.10 %) | 59 (21.38 %) | 120 (43.48 %) |
| Negative | 77 (27.90 %) | 79 (28.62 %) | 156 (56.52 %) |
| **HER2 Status** |  |  |  |
| Positive | 82 (29.71 %) | 78 (28.26 %) | 160 (57.97 %) |
| Negative | 56 (20.29 %) | 60 (21.74 %) | 116 (42.03 %) |

**Supplementary Table 3.** PARTNER.

|  | Non-pCR<br>(N=118) | pCR<br>(N=165) | Overall<br>(N=283) |
| --- | --- | --- | --- |
| **TRIGS.TIL.density** |  |  |  |
| Mean (SD) | -6.53 (0.661) | -6.45 (0.667) | -6.48 (0.665) |
| Median [Min, Max] | -6.42 [-8.27, -4.97] | -6.41 [-8.85, -4.99] | -6.41 [-8.85, -4.97] |
| **HoverNet.TIL.density** |  |  |  |
| Mean (SD) | -6.20 (0.981) | -5.89 (0.844) | -6.02 (0.915) |
| Median [Min, Max] | -5.94 [-9.48, -4.12] | -5.80 [-8.27, -4.39] | -5.90 [-9.48, -4.12] |
| Missing | 0 (0%) | 1 (0.6%) | 1 (0.4%) |
| **MuTILs.TIL.density** |  |  |  |
| Mean (SD) | -6.87 (1.19) | -6.46 (1.03) | -6.63 (1.11) |
| Median [Min, Max] | -6.79 [-9.79, -4.45] | -6.39 [-9.01, -4.54] | -6.50 [-9.79, -4.45] |
| Missing | 27 (22.9%) | 38 (23.0%) | 65 (23.0%) |
| **SAM.TIL.density** |  |  |  |
| Mean (SD) | -6.62 (0.784) | -6.40 (0.750) | -6.49 (0.770) |
| Median [Min, Max] | -6.49 [-8.58, -4.65] | -6.35 [-9.36, -4.88] | -6.38 [-9.36, -4.65] |
| **Tumour Infiltrating Lymphocytes score** |  |  |  |
| Mean (SD) | 28.4 (24.6) | 36.6 (27.3) | 33.1 (26.5) |
| Median [Min, Max] | 20.0 [1.00, 90.0] | 30.0 [0, 100] | 30.0 [0, 100] |
| **Tumour size** |  |  |  |
| =< 50mm | 112 (94.9%) | 159 (96.4%) | 271 (95.8%) |
| > 50mm | 6 (5.1%) | 6 (3.6%) | 12 (4.2%) |
| **Cancer type** |  |  |  |
| gBRCAwt | 108 (91.5%) | 137 (83.0%) | 245 (86.6%) |
| BRCA | 10 (8.5%) | 28 (17.0%) | 38 (13.4%) |
| **Histopathological involvement of axillary nodes** |  |  |  |
| No | 78 (66.1%) | 119 (72.1%) | 197 (69.6%) |
| Yes | 40 (33.9%) | 46 (27.9%) | 86 (30.4%) |

